# Comparison of viral levels in individuals with or without symptoms at time of COVID-19 testing among 32,480 residents and staff of nursing homes and assisted living facilities in Massachusetts

**DOI:** 10.1101/2020.07.20.20157792

**Authors:** Niall J. Lennon, Roby P. Bhattacharyya, Michael J. Mina, Heidi L. Rehm, Deborah T. Hung, Sandra Smole, Ann Woolley, Eric S. Lander, Stacey B. Gabriel

## Abstract

**Background:** Transmission of COVID-19 from people without symptoms poses considerable challenges to public health containment measures. The distribution of viral loads in individuals with and without symptoms remains uncertain. Comprehensive cross-sectional screening of all individuals in a given setting provides an unbiased way to assess viral loads independent of symptoms, which informs transmission risks. COVID-19 cases initially peaked in Massachusetts in mid-April 2020 before declining through June, and congregate living facilities were particularly affected during this early surge. We performed a retrospective analysis of data from a large public health-directed outbreak response initiative that involved comprehensive screening within nursing homes and assisted living facilities in Massachusetts to compare nasopharyngeal (NP) viral loads (as measured by RT-PCR cycle threshold (Ct) levels) in residents and staff to inform our ability to detect SARS-CoV-2 in individuals with or without symptoms in the population.

**Methods:** Between April 9 and June 9, 2020, we tested NP swabs from 32,480 unique individuals comprising staff and residents of the majority of nursing homes and assisted living facilities in Massachusetts. Under the direction of the MA Department of Public Health (MDPH), symptomatology at the time of sampling and demographic information was provided by each facility for each individual to facilitate reporting to health officials. NP swabs were collected, RNA extracted, and SARS-CoV-2 testing performed using quantitative reverse-transcriptase polymerase chain reaction (qRT-PCR).

**Results:** The nursing home and assisted living facilities resident cohort (N =16,966) was 65% female with a mean age of 82 years (SD 13 yrs). The staff cohort (N = 15,514) was 76% female with a median age of 45 (SD 15 yrs). A total 2654 residents (15.5%) and 624 staff (4.1%) tested positive for SARS-CoV-2. 12.7% of residents and 3.7% of staff without symptoms tested positive for SARS-CoV-2, compared to 53.1% of residents and 18.2% of staff with symptoms. Of the individuals who tested positive, 70.8% of residents and 92.4% of staff lacked symptoms at the time of testing. In aggregate, the distributions of Cts for viral probes used in the qRT-PCR assay were very similar, with a statistically but not meaningfully different mean (ΔCt 0.71 cycles, p = 0.006) and a similar range (12-38 cycles), between populations with and without symptoms over the entire time period, across all sub-categories examined (age, race, ethnicity, sex, resident/staff). Importantly, the Ct mean values and range were indistinguishable between the populations by symptom class during the peak of the outbreak in Massachusetts, with a Ct gap appearing only later in the survey period, reaching >3 cycles (p ≤ 0.001) for facilities sampled during the last two weeks of the study.

**Conclusions:** In a large cohort of individuals screened for SARS-CoV-2 by qRT-PCR, we found strikingly similar distributions of viral load in patients with or without symptoms at the time of testing during the local peak of the epidemic; as the epidemic waned, individuals without symptoms at the time of testing had lower viral loads. The size of the study population, including both staff and residents spanning a wide range of ages, provides a comprehensive cross-sectional point prevalence measurement of viral burden in a study spanning 2 months. Because the distributions of viral loads in infected individuals irrespective of symptomatology are very similar, existing testing modalities that have been validated for detection of SARS-CoV-2 RNA in symptomatic patients should perform similarly in individuals without symptoms at the time of testing.

## Background

Despite the public health importance of coronavirus infectious disease 2019 (COVID-19), the relationship between viral load, symptom severity, and transmission risk remain poorly understood. As the primary focus on controlling community transmission of Severe Acute Respiratory Syndrome Coronavirus 2 (SARS-CoV-2) expands from existing outbreak response to new outbreak surveillance, it is increasingly important to be able to perform accurate viral testing in individuals that do not show COVID-19 symptoms at the time of testing^1,2,3,4,5,6,7^. While still evolving, recent reports suggest that a substantial fraction of SARS-CoV-2 spread occurs from such infected individuals without symptoms at the time of transmission^8,9^. Detecting such individuals before they expose others to the virus could therefore play an important role in limiting spread within a population.

To determine whether current testing methods are appropriate for testing individuals without symptoms, it is necessary to understand the relative distributions of viral loads in patients with and without symptoms. (We employ the commonly-used operational definition of viral load based on the quantity of viral RNA in a patient specimen as measured by qRT-PCR; we address limitations of this measure in the Discussion.) A major question with testing individuals without symptoms at the time of sampling is whether their viral loads will be substantially lower than in symptomatic individuals, as has been demonstrated for influenza^10^, so as to compromise the reliability of existing assays for detecting virus in these infected individuals. Assays to detect the presence of SARS-CoV-2 RNA in samples often have limits of detection between several hundred to several thousand viral genomes per milliliter (mL)^11^.

To date, viral loads in individuals without symptoms have not been extensively studied because testing has been primarily focused on individuals with symptoms^12,13^. In contrast to influenza, where asymptomatic or paucisymptomatic individuals have been reported to have 10 to 100-fold less virus than symptomatic individuals^10^, several recent small studies have found similar SARS-CoV-2 RNA levels in infected individuals irrespective of symptoms. In one study of 30 individuals in quarantine, 13 asymptomatic individuals had the same viral loads as 17 symptomatic patients at baseline^14^. A second study of 37 hospitalized asymptomatic individuals also found similar viral loads as their symptomatic counterparts^15^. Among those who do show symptoms severe enough to require hospitalization, one recent study showed that initial viral load on admission was associated with increased risk for death or intubation^12^. However, given the wide range of viral load over time within each patient^811^, higher viral load at the time of admission could simply be a proxy for those who became sick earlier in the course of their infection. Given the small number of asymptomatic patients in these studies, it is important to study viral load data by symptom status across a larger population, including across age, sex, baseline health status, and other factors such as comorbidities.

To understand the biological relationship between symptomatology and viral loads, certain study designs are preferable. Systematic screens of all individuals in a given setting, without regard to the presence of symptoms, are preferable to studies of individuals who present for testing or for whom testing is ordered: the latter design is likely to involve significant selection bias, because most people without symptoms are not currently tested, and those who are tested are unlikely to be representative of the whole. Studies occurring relatively early in an epidemic are also preferable to studies at later times, because the infection dates are likely to be more closely synchronized.

As the local epidemic neared its peak in April 2020, in response to several large outbreaks, the Commonwealth of Massachusetts initiated an aggressive, systematic program to perform comprehensive viral testing of all staff and residents in all skilled nursing facilities and assisted living facilities, regardless of whether individuals showed COVID-19 symptoms. For the majority of these facilities, specimens (collected by nasopharyngeal swabs) were sent to either the Massachusetts Department of Public Health State Public Health Laboratory (MASPHL) or its contracted reference laboratory, the Clinical Research Sequencing Platform (CRSP) at the Broad Institute for viral testing via a real-time qPCR assay. Between April 9 and June 9, 2020, the Broad Institute laboratory performed 32,480 unique individual diagnostic tests on people who were identified as residents or staff at 366 skilled nursing facilities and assisted living facilities in Massachusetts. During the period of this study, overall COVID-19 burden in the state peaked at >3000 confirmed cases per day on April 17 (week 2 of the study) and declined thereafter, dropping over 7-fold by the end of the study period (Figure S1)^16^. For each individual, the facility reported symptomatology at the time of sampling as ascertained by the onsite physician or nursing staff, as well as basic demographic information to facilitate reporting to the Massachusetts Department of Public Health.

These data (symptomatic status reported by the facility, demographic data, and viral load measured by the RT-PCR assay) provide a large point-prevalence survey. Because the vast majority of the individuals were sampled only once and longitudinal history was not available, these data do not distinguish individuals who were durably asymptomatic from those who subsequently developed symptoms and were thus presymptomatic at the time of testing. We therefore refer to these people throughout as individuals without symptoms at the time of testing, to clarify that we do not attempt to distinguish asymptomatic from presymptomatic infection. While these two groups have different implications for contact-tracing efforts, either may transmit disease in the absence of symptoms^18^ and are thus crucial to study quantitatively.

The primary issue addressed in this paper is the comparison of estimated viral load distributions (as measured by cycle threshold (Ct) for viral detection in the real-time RT-PCR assay) from nasopharyngeal (NP) swabs between individuals with and without symptoms at the time of testing. The Ct value measures the number of amplification cycles required to detect cDNA produced from viral RNA; a higher Ct value indicates less viral RNA in the sample. While a small, statistically significant difference between the populations could be detected over the entire study period, notably, no difference was detected at the peak of the epidemic, with a small gap between their mean Ct values emerging as the epidemic waned locally. The results suggest that the distribution of viral load in infected individuals with or without symptoms at the time of testing is similar, and thus assays that reliably detect virus in symptomatic individuals should perform equally well for individuals without symptoms.

## Methods

### Study Population

Between April 9 and June 9, 2020, the Broad Institute’s CLIA-certified clinical laboratory received NP swab specimens for SARS-CoV-2 testing from 366 skilled nursing facilities, nursing homes, and assisted living facilities across the Commonwealth of Massachusetts. Determination of the population selected for testing was based on the CDC 2019-Novel Coronavirus (2019-nCoV) Real-Time RT-PCR Diagnostic Panel (CDC-006-00019, Rev 2) instructions for use in which “2019-nCoV testing may be indicated as part of a public health investigation”. Swabs were collected by trained staff onsite at the homes (a minority of samples) or by the Massachusetts National Guard (MANG) (the majority of samples). To facilitate collection, MANG deployed twelve medical teams each consisting of medics, decontamination personnel, a non-commissioned officer in charge, and other support members of the MANG. Eligibility criteria for testing were broad: the intention was to test every resident and every staff member of every facility. Testing was performed on 32,480 unique individuals, with a small proportion (6.7%) tested more than once during the period. For individuals tested more than once, only data from the first test are reported here in order to avoid duplication.

### Symptom and Demographic Information

Beginning in the second week of the testing program (labeled as Week 2, April 17-23), the facility filled out a requisition form for each individual swabbed that asked whether the individual did or did not show COVID-19 symptoms. While the requisition form did not request specific details about the types or severity of symptoms and thus assessments may not be completely uniform across facilities, a binary judgment of symptomatology was made by the facility’s trained skilled nursing staff or physician on-site. Longitudinal information was not available about whether individuals without symptoms at the time of collection previously had or later developed symptoms. However, because most nursing facilities in Massachusetts during the study period required negative SARS-CoV-2 RT-PCR before accepting patients with known or suspected COVID-19 in transfer from acute care hospitals, those testing positive would not include many post-symptomatic, persistently positive individuals convalescing from known COVID-19.

For each individual swabbed, the requisition form also requested personal and demographic information consisting of name, date of birth, race, ethnicity, sex, symptom status (symptomatic or asymptomatic), and whether they were a resident or a staff member. In the minority of responses where answers on the requisition form were blank or unclear, data were coded as missing (see Table 1).

**Table 1.** Numbers of individual participants in each of the categories for demographic variables collected on the diagnostic test requisition form. % of the category within each participant type (Resident or Staff) is also shown. Individual participant ages were grouped into decade of life to preserve anonymity. Test results for each participant type category is also shown. Positive indicates the detection of SARS-CoV-2 in the specimen. Negative indicates no detectable SARS-CoV-2 in the specimen. Inconclusive indicates a case where one viral probe (N1 or N2) is positive but the other is negative. The human RP probe must be positive for a specimen with negative viral probes to be called negative. Otherwise that test would be called invalid.

|  | **Resident (16,966)** |  | **Staff (15,514)** |  |
| --- | --- | --- | --- | --- |
| **Participant Gender** | Number | % of Resident | Number | % of Staff |
| Female | 11040 | 65.1% | 11763 | 75.8% |
| Male | 5109 | 30.1% | 3051 | 19.7% |
| Other | 4 | 0.02% | 6 | 0.04% |
| Missing | 813 | 4.8% | 694 | 4.5% |
| **Participant Race** |  |  |  |  |
| American Indian or Alaska Native | 147 | 0.9% | 58 | 0.3% |
| Asian | 274 | 1.6% | 461 | 2.7% |
| Black or African-American | 751 | 4.4% | 3605 | 21.2% |
| Native Hawaiian or Other Pacific Islander | 8 | 0.0% | 19 | 0.1% |
| Other | 277 | 1.6% | 1035 | 6.1% |
| White | 14369 | 84.7% | 8454 | 49.8% |
| Missing | 1140 | 6.7% | 1882 | 11.1% |
| **Participant Ethnicity** |  |  |  |  |
| Hispanic or Latino | 357 | 2.1% | 1724 | 10.2% |
| Not Hispanic or Latino | 12776 | 75.3% | 9724 | 57.3% |
| Missing | 3833 | 22.6% | 4066 | 24.0% |
| **Decade of Life** |  |  |  |  |
| 10-19 | 5 | 0.03% | 751 | 4.4% |
| 20-29 | 56 | 0.3% | 2505 | 14.8% |
| 30-39 | 88 | 0.5% | 2609 | 15.4% |
| 40-49 | 188 | 1.1% | 3093 | 18.2% |
| 50-59 | 686 | 4.0% | 3704 | 21.8% |
| 60-69 | 1733 | 10.2% | 2358 | 13.9% |
| 70-79 | 3270 | 19.3% | 365 | 2.2% |
| 80-89 | 5857 | 34.5% | 35 | 0.2% |
| 90-99 | 4778 | 28.2% | 18 | 0.1% |
| 100-115 | 263 | 1.6% | 2 | 0.01% |
| Missing | 42 | 0.2% | 74 | 0.4% |
| **Symptomatology** |  |  |  |  |
| No symptoms | 13341 | 78.6% | 13258 | 78.1% |
| Symptomatic | 1316 | 7.8% | 220 | 1.3% |
| Missing | 2309 | 13.6% | 2036 | 12.0% |
| **Test Result** |  |  |  |  |
| Positive | 2654 | 15.5% | 624 | 4.1% |
| Negative | 14196 | 83.9% | 14833 | 95.6% |
| Inconclusive | 116 | 0.7% | 57 | 0.4% |

### Laboratory Testing

NP swabs were placed in barcoded tubes with 3 mL of viral transport medium (VTM), transported in coolers with ice packs, and delivered to the laboratory on the day of collection. Symptomatology and demographic information listed on the test requisition form were entered into a database and associated with the barcode on the specimen tube. RNA was extracted from 50ul of VTM using the MagMax-96 RNA extraction kit (Thermo Fisher) on a Bravo liquid handler platform (Agilent). One-step real-time reverse transcriptase–polymerase chain reaction (RT-PCR) was performed on a QuantStudio 7 (Applied Biosystems), using a laboratory developed SARS-CoV2 CDC assay protocol run under the FDA’s Emergency Use Authorization framework; cycle threshold (Ct) values were reported for two viral probes, the N1 and N2 viral nucleocapsid protein gene regions, and a RNaseP human gene control (RP)^17^). Ct values lower than 40 cycles for both N1 and N2 indicate a diagnostic qualitative positive result for SARS-CoV-2 (a single positive viral probe was reported as Inconclusive). Viral loads (copies/mL) were estimated by interpolation from a standard curve generated by serial dilutions of a synthetic RNA construct (Twist Biosciences, CA) containing the viral N2 target sequence; the Ct values correlated strongly with the logarithm of RNA concentration (R^2^ > 0.99), with the observed range from Ct =12 cycles to Ct = 38 cycles corresponding to viral loads ranging from ∼1.9 billion copies/mL to 8 copies/mL, respectively.

### Analyses

The distribution of Ct values were plotted as a function of various metadata. For simplicity of analysis and presentation the Ct values for the N1 and N2 probes in positive patients were averaged. One-way ANOVA and pooled t-tests were performed between subpopulations to determine the significance of differences in Ct values. All analyses were completed with SAS JMP software, version 13 (SAS Institute). Internal Ct data were collected as part of the diagnostic efforts as part of this public health response and were deemed exempt human subjects research by the Broad Institute Office of Research Subject Protection and approved with waiver of informed consent by the MA Department of Public Health’s Institutional Review Board.

## Results

### Summary

Across all facilities, 2654 residents (15.5%) and 624 staff (4.1%) tested positive for SARS-CoV-2. Among the residents, 78.6% were listed as asymptomatic at the time of swabbing and 7.8% were listed as symptomatic (remaining resident forms did not indicate symptomatology). Among the staff, 78.1% were listed as asymptomatic at the time of swabbing, and 1.3% were listed as symptomatic (remaining staff forms did not indicate symptomatology). Mean age of residents was 82 years (SD 13, range 17 to 114), while mean age of staff was 45 years (SD 15, range 16 to 101). Table 1 shows the demographics and aggregate results for the resident and staff cohorts.

### Positive rates by symptom class

Among 13,341 residents who lacked symptoms at the time of swabbing, 1692 (12.7%) were positive, compared with 487 (3.7%) of 12,724 staff without symptoms. 699 (53.1%) of 1316 residents with symptoms tested positive, compared with 40 (18.2%) of 220 staff with symptoms (Table 2a).

**Table 2.** Test results in Residents and Staff are shown as a function of symptomatology (2a). Resident and staff symptomatology is also shown as a function of the test result (2b).

| **Symptomatology** | **Test Result** | **Resident** |  | **Staff** |  |
| --- | --- | --- | --- | --- | --- |
|  |  | **Number** | **%** | **Number** | **%** |
| No symptoms | Positive | 1692 | 12.7% | 487 | 3.7% |
|  | Negative | 11564 | 86.7% | 12724 | 96.0% |
|  | Inconclusive | 85 | 0.6% | 47 | 0.4% |
| Symptomatic | Positive | 699 | 53.1% | 40 | 18.2% |
|  | Negative | 597 | 45.4% | 180 | 81.8% |
|  | Inconclusive | 20 | 1.5% | 0 | 0.0% |

| **Test Result** | **Symptomatology** | **Resident** |  | **Staff** |  |
| --- | --- | --- | --- | --- | --- |
|  |  | **Number** | **%** | **Number** | **%** |
| Positive | No symptoms | 1692 | 70.8% | 487 | 92.4% |
|  | Symptomatic | 699 | 29.2% | 40 | 7.6% |
| Negative | No symptoms | 11564 | 95.1% | 12724 | 98.6% |
|  | Symptomatic | 597 | 4.9% | 180 | 1.4% |
| Inconclusive | No symptoms | 85 | 81.0% | 47 | 100.0% |
|  | Symptomatic | 20 | 19.0% | 0 | 0.0% |

### Symptomatology by test results

Of the individuals who tested positive, a substantial majority lacked symptoms at the time of sampling, including 1692 (70.8%) of 2391 residents and 487 (92.4%) of 527 staff (Table 2b).

### Comparison of viral load between individuals with and without symptoms at the time of testing over the entire 6 week study period

Among individuals who tested positive, over the entire time period, the Ct levels for viral load (as an average of the N1 and N2 probes) covered a broad range, from 11.6 to 37.7 cycles in individuals without symptoms and 11.9 to 37 cycles in individuals with symptoms (Figure 1a,b), while the Ct for the human host probe (RP) was more tightly distributed around a mean of 28.9 (SD 2.4) and 28.1 (SD 2.7) cycles for each population (Figure 1c,d).

**Figure 1.**
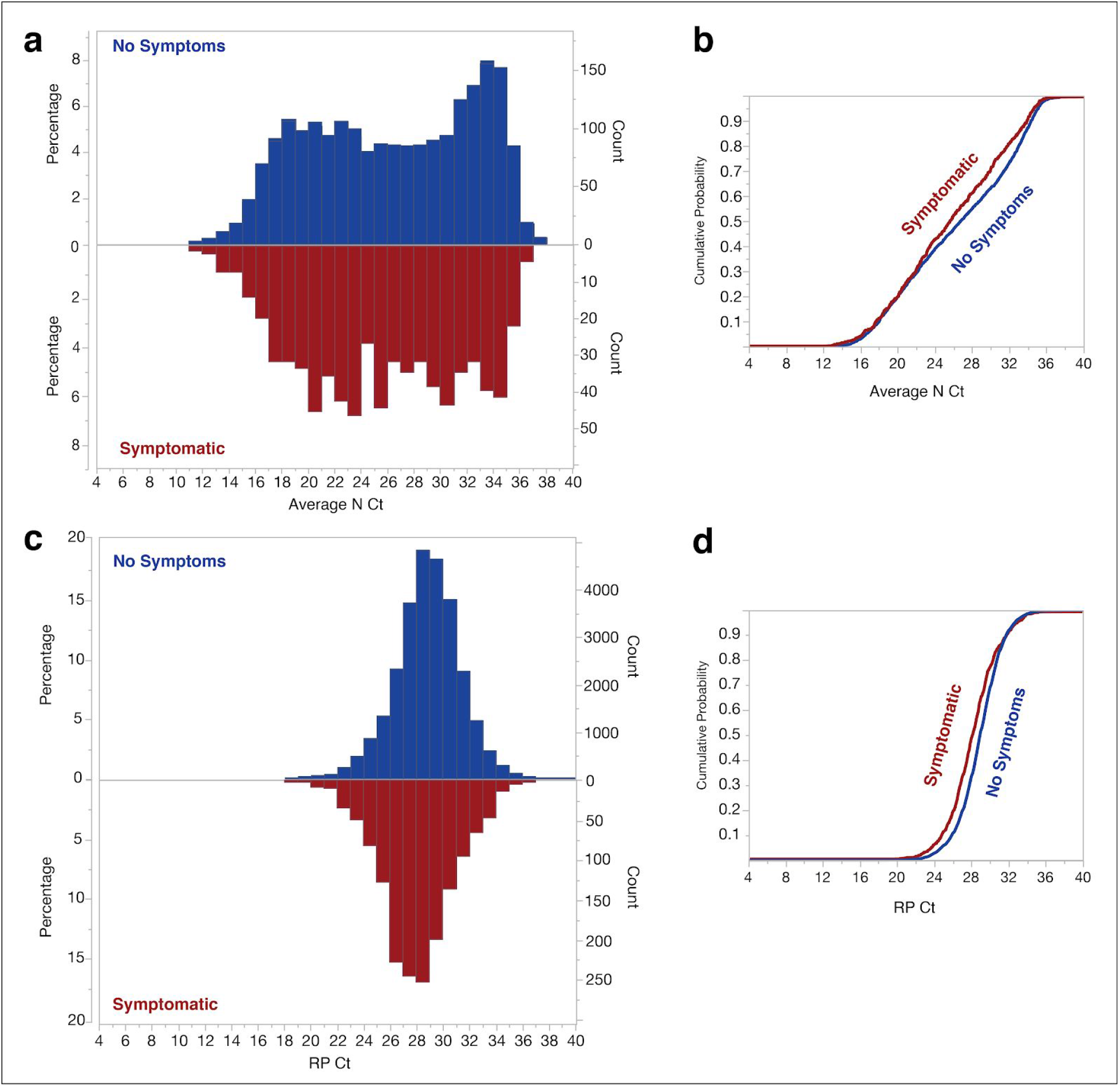
Cumulative and actual distribution of Cts by Symptom class across all unique individuals tested. Histograms of Cts for the N and RP probes are shown in (a) and (c) respectively, with each bar indicating samples with values between the tick marks. No symptom distribution (blue) is shown above the line with the symptomatic distribution (red) below the line in both cases. Panels b and d indicates the cumulative distribution of the N probes (averaged across N1 and N2) (b) and the RP probe (d) colored by symptom class (Red = symptomatic; Blue = No symptoms).

The distributions for the viral level differed slightly between individuals with and without symptoms, with a difference in mean Ct of only 0.71 cycles (26.4 vs 25.7, p=0.006) and a slightly higher proportion of individuals with Ct ≥ 30 cycles (36% for individuals without symptoms vs. 29.2% for those with symptoms) (Figure 1a,b). Similarly, the mean Ct for the human host probe differed by 0.74 cycles (p=0.0001) between these two populations (Figure 1c,d).

Despite the statistically detectable differences, both individuals with and without symptoms show substantially similar distributions down to the limit of detection of the assay, with only a small difference in mean Ct value. For context, test developers and the FDA typically use a Ct difference of <3 cycles as an indicator of substantial equivalence between viral testing methods. Furthermore, the observed differences in Ct are less than the typical variability in sampling efficiency, as reflected in the RP probe Ct distributions (SD 2.4 and 2.6 cycles).

### Cumulative distributions of virus abundance

The ∼250 million-fold range of viral loads observed (from Ct of 12 to 38 for the N2 probe) is consistent with prior studies^18,19,20,21^. This wide range implies that the vast majority of total viral load in the population sampled was carried by a minority of individuals with the lowest Ct values. Following an analysis in a recent report^18^, we calculated the proportion of total viral load carried by those individuals with the highest x% of viral load, for each value of x, by weighting the number of people in each bin of Ct values by the viral load corresponding to each bin of Ct values (Figure 2). For individuals without symptoms, 9% of individuals (those with Ct ≤ 17 cycles) harbored 90% of the total virus in the population sampled, and 24% of individuals (those with Ct ≤ 20 cycles) harbored 99%. Similar values were seen for the set of individuals with symptoms (Figure 2, inset).

**Figure 2.**
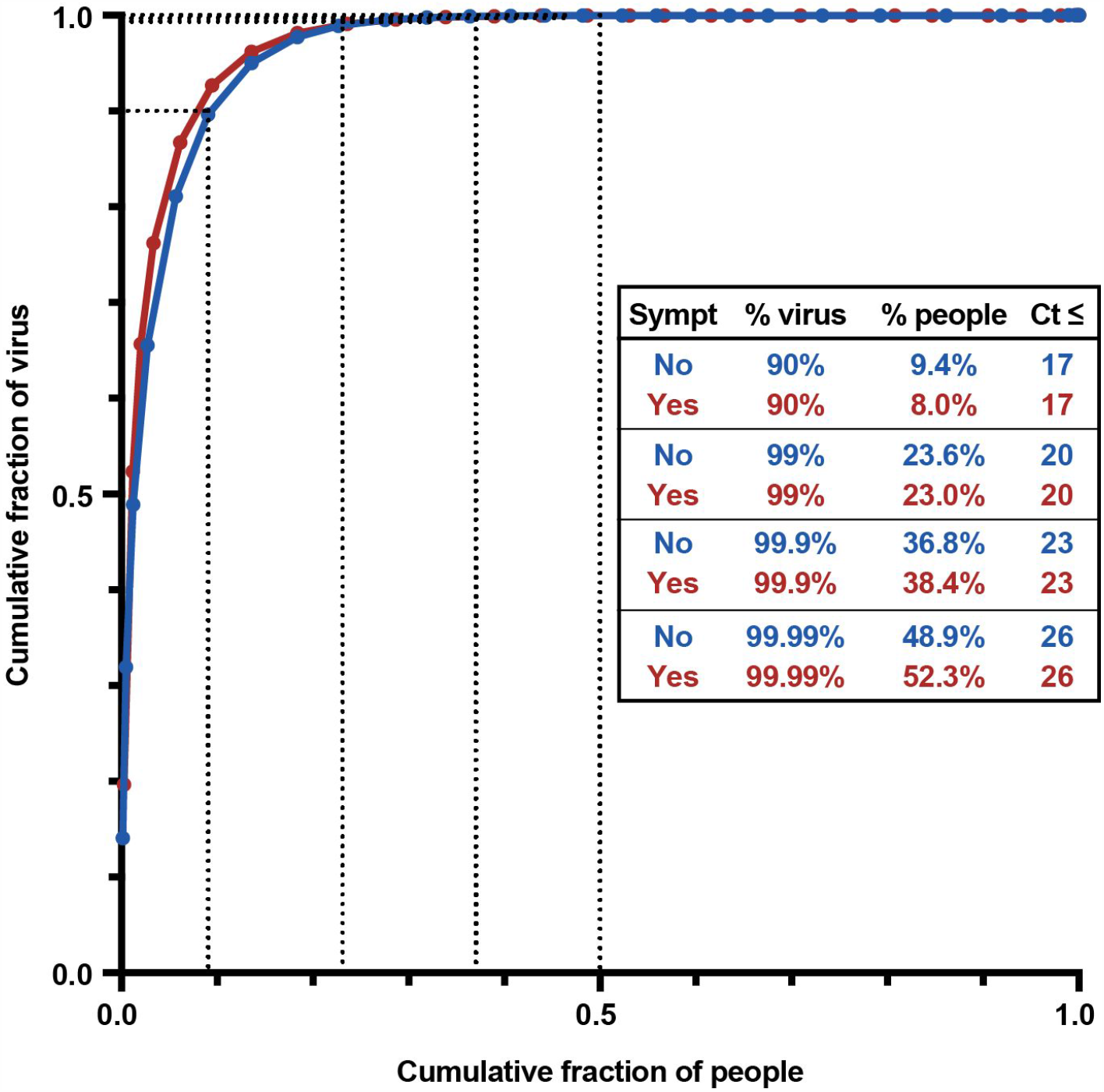
Cumulative distribution of total viral load across individuals (red, individuals with symptoms; blue, individuals without symptoms). The fraction of total viral load (y-axis) harbored by individuals with a given Ct value was calculated by multiplying the number of individuals with each Ct value by the viral load corresponding to that Ct value, and then normalizing these numbers to sum to 1. These fractions were then used to create the cumulative distribution plot, with people ordered from highest to lowest viral load (lowest to highest Ct value) along the x-axis. Dotted lines correspond to 90%, 99%, 99.9%, and 99.99% of cumulative viral burden, with the corresponding percentage of individuals tabulated in the inset, along with the corresponding Ct threshold.

### Variation of viral loads over time

When the distribution of viral loads between individuals with and without symptoms was compared over time, on a weekly basis, no difference was observed between the two populations, either in mean Ct value or range, during the time period that coincided with the peak outbreak of COVID-19 in Massachusetts (April 17-23) (Figure 3; Figure S1). However, with time, a gap emerged, with mean Ct value shifting higher in the population without symptoms while remaining essentially unchanged for symptomatic patients throughout the testing period. Specifically, individuals without symptoms tested in the last two weeks of this study during weeks 5 and 6 (May 7-20) had Ct values >3 cycles greater (less virus) than symptomatic individuals (p = 0.0013 and 0.0007 for weeks 5 and 6, respectively). These weeks corresponded to a waning of the epidemic in Massachusetts, as weekly case counts declined by 7-fold from the beginning to the end of the study period.

**Figure 3.**
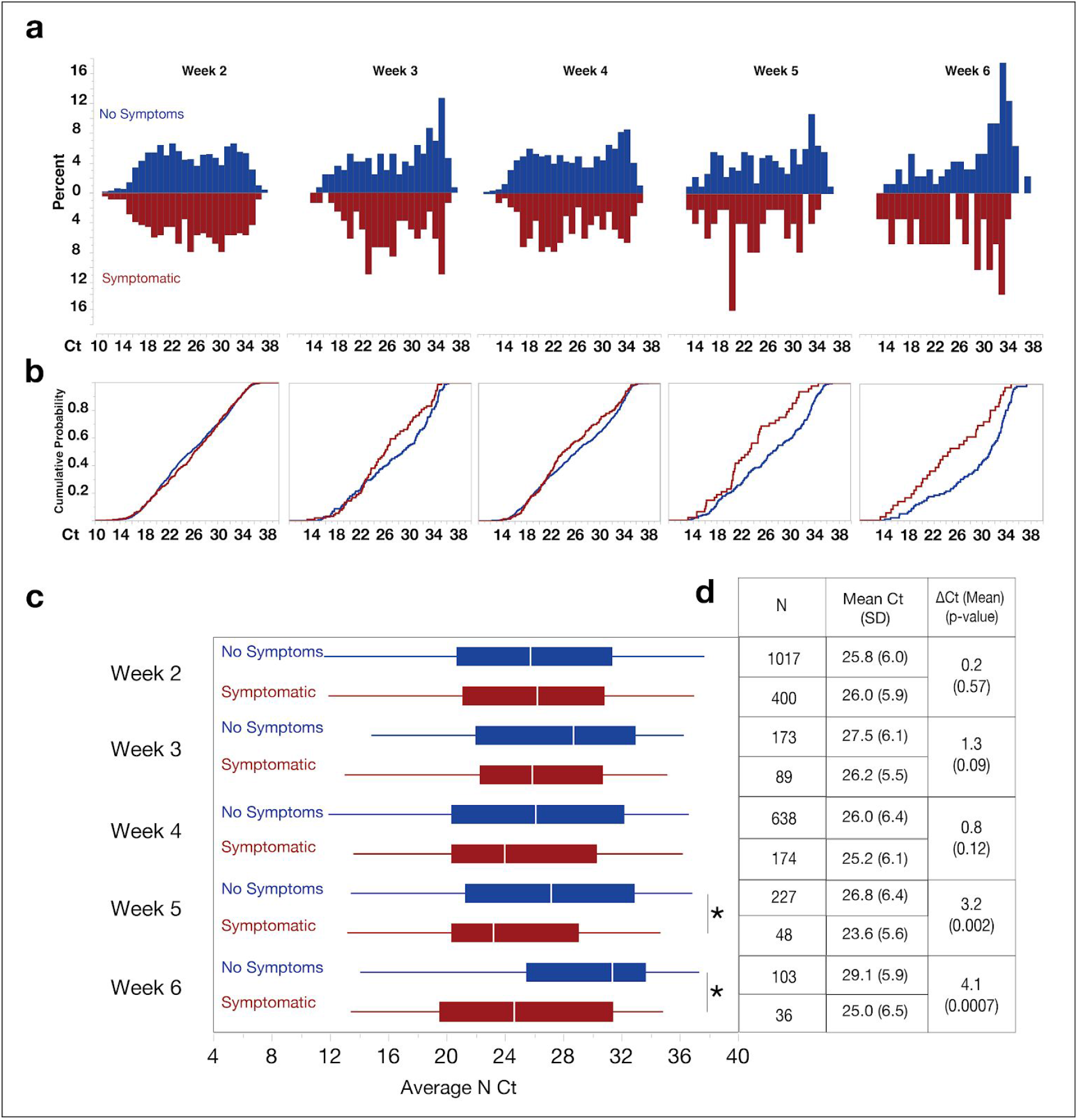
(a) Distribution of Cts by Symptom class across unique individuals tested by week of study (Week 2, the first week in which symptom data were collected, spans April 17-23, 2020; Week 3 spans April 24-30; and so on) by symptom status (Blue = no symptoms at time of testing, Red = symptoms at time of testing). Weeks with fewer than 20 data points in either category are not shown. Week 1 data is not shown as symptom class was not captured in the first week of testing. (b) Cumulative distribution plots of the data from (a) overlaid (Blue = no symptoms at time of testing, Red = symptoms at time of testing) (c) Box-plots of the average viral N probe (N1 and N2) Ct by week and symptom class, with vertical line at median, colored boxes at IQR, and whiskers showing full range. Asterisks indicate statistically significant differences within a sub-category. Table of sample size and mean Ct with standard deviation (SD) is shown in (c). Also shown is the sub-category ΔCt between the symptomatic and no symptom cases and the associated p-value.

### Effect of age and other demographic variables

Since age dramatically affects COVID-19 severity^22^, to examine whether age modified the relationship between viral load and symptom class, we partitioned individuals by decade of life and assessed mean viral level in the resident and staff cohorts separately (Table 3). We observed that symptomatology does not significantly alter viral level in any of the age groups over the entire study period, regardless of whether the person is a staff member or a resident. We also looked at the percentage of positive cases by symptom class across different age groups for both residents and staff (Figure 4). Notably, in each age group, the majority of positive results occurred in persons listed as having no symptoms at the time of swabbing.

Other available demographic variables (sex, race, ethnicity, resident vs staff) were examined to see if they modified the relationship between viral load between individuals with and without symptoms at the time of testing (Figure 5), since social determinants of health^2324^ and baseline health status^22^ also impact COVID-19 outcomes. Again, statistically significant but numerically small differences were observed between those with and without symptoms in some categories (ΔCt = 0.75 cycles overall, range 0.8 - 1.2 cycle difference among demographic classes with p < 0.05).

**Table 3.**
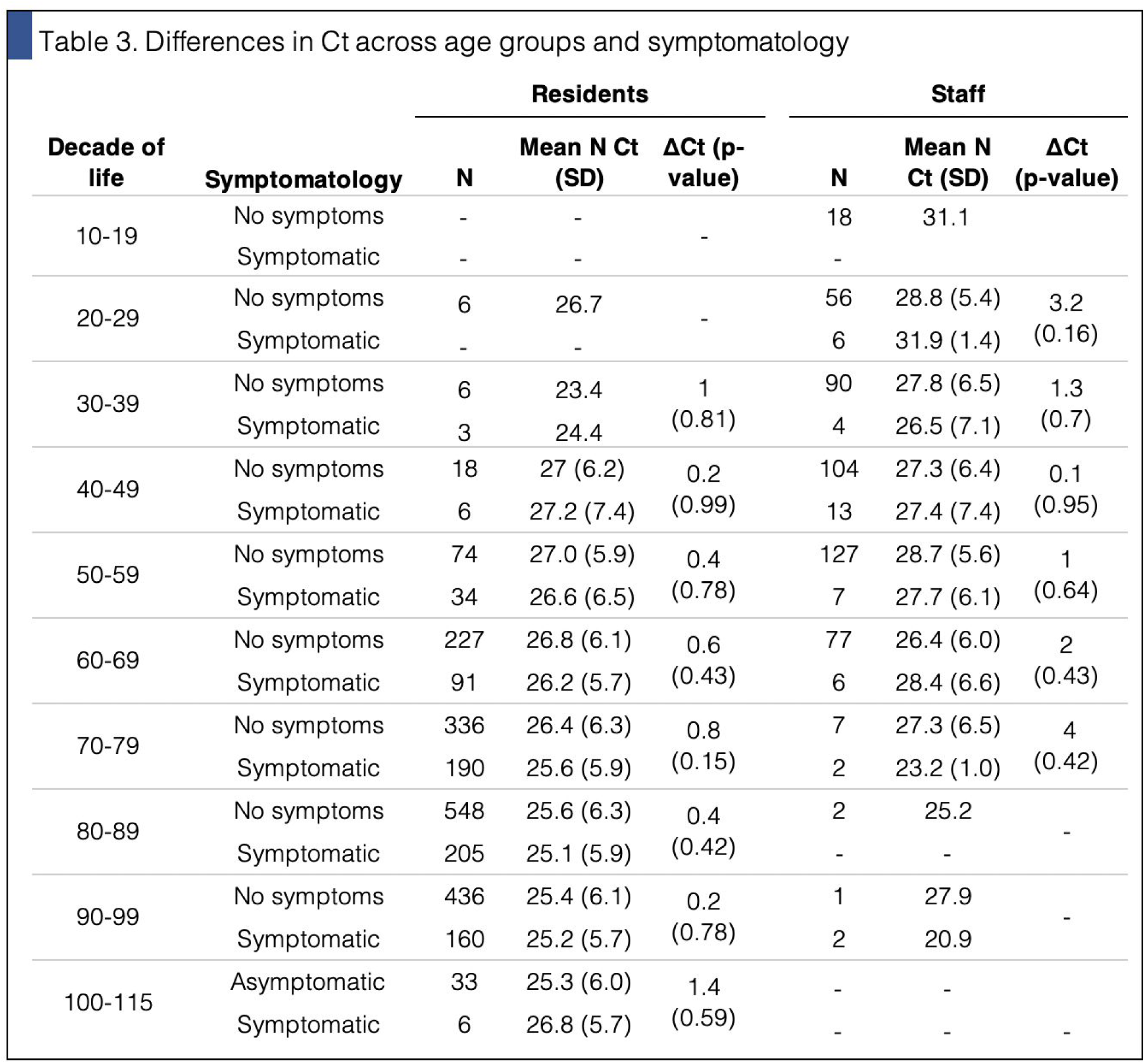
Examination of the mean N probe (average of N1 and N2) Ct by age group across residents and staff and symptom class. SD: standard deviation.

**Figure 4.**
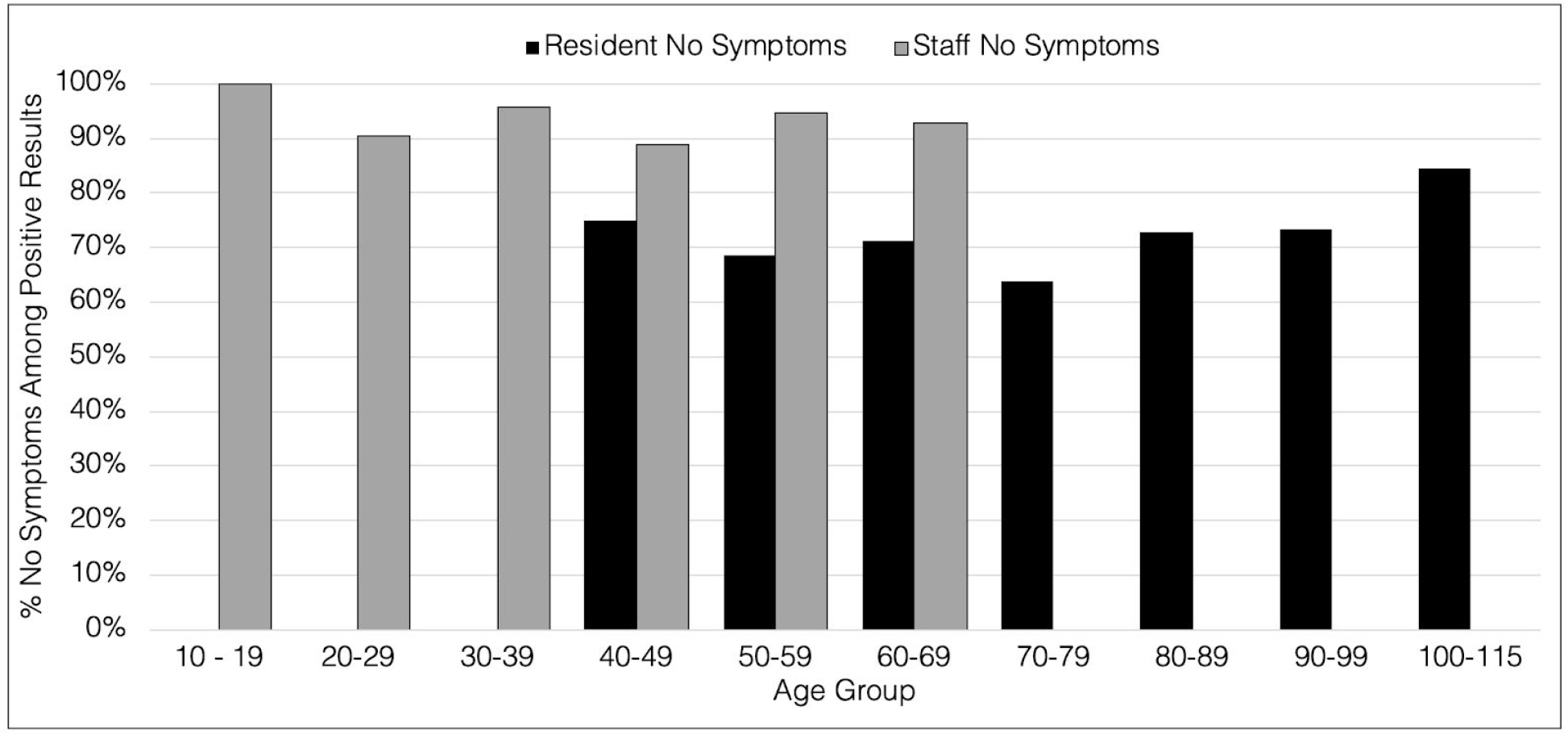
Percentage of the positive cases within the staff and resident groups that had no symptom at each age group level.

**Figure 5.**
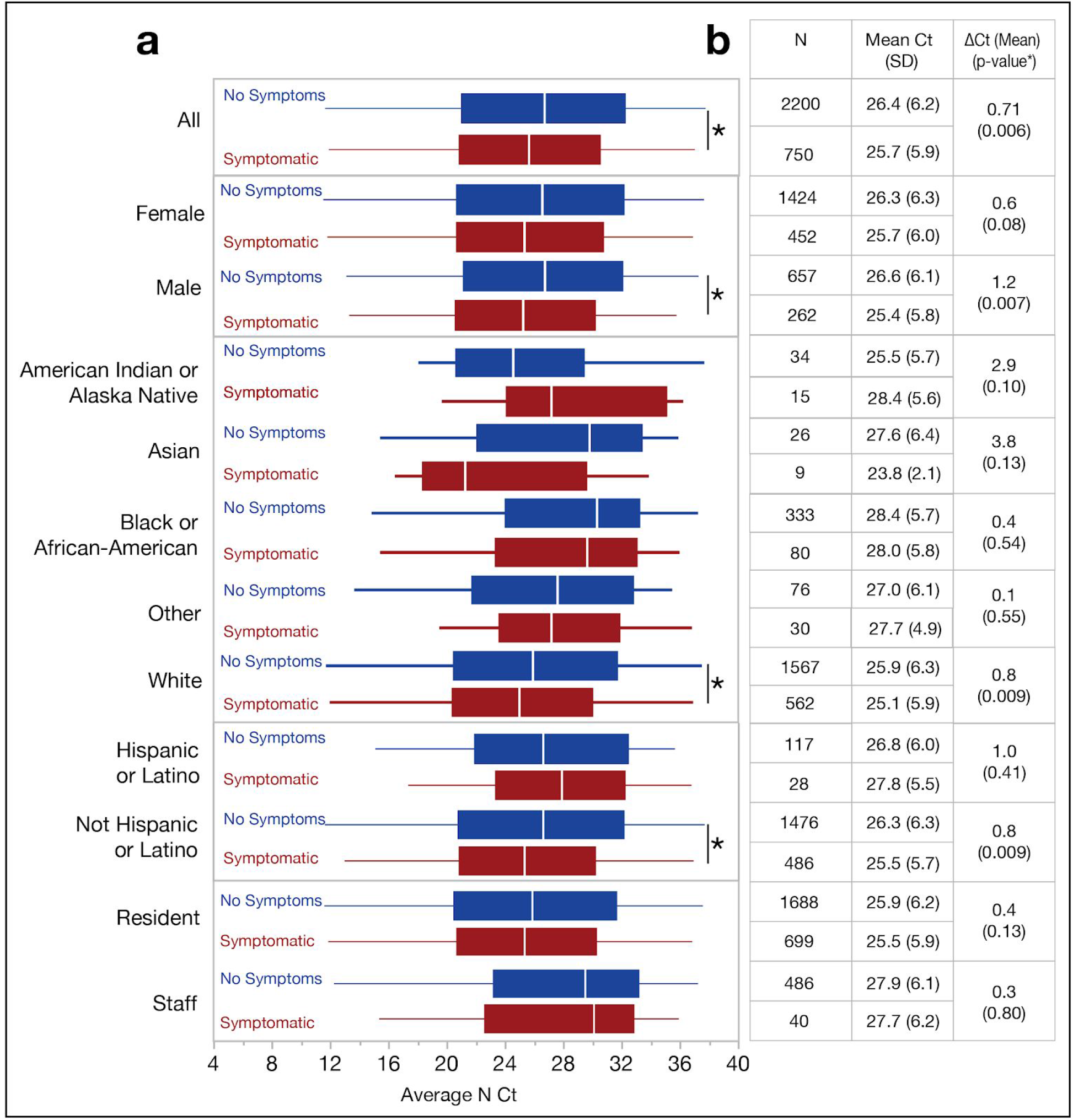
Ct distributions across demographic and symptom categories. Any sub-category with <20 data points was removed. Box-plots of the average viral N probe (N1 and N2) Ct by category and symptom class are shown in (a), with vertical line at median, colored boxes at IQR, and whiskers showing full range. Asterisks indicate statistically significant differences within a sub-category. Table of sample size and mean Ct with standard deviation (SD) is shown in (b). Also shown is the sub-category ΔCt between the symptomatic and no symptom cases and the associated p-value.

## Discussion

COVID-19 has become a devastating pandemic because of its considerable morbidity and mortality^25^ coupled to its frequent spread from individuals who do not show symptoms at the time of transmission ^8,25^. Control measures that aim to detect a substantial portion of transmission events thus require diagnosis of infected patients who do not display symptoms at the time of testing^26^. Prior reports of large-scale cross-sectional SARS-CoV-2 testing have not reported quantitative viral load in individuals without symptoms at the time of testing

By comprehensively screening 16,966 residents and 15,514 staff of residential nursing facilities in Massachusetts, while capturing simultaneous symptom classification from healthcare providers, we generated quantitative RT-PCR data from 2179 and 739 subjects without and with symptoms, respectively, the largest cohort of individuals without symptoms at the time of testing reported to date. The Ct distributions between the two populations over the entire time period were remarkably similar. They were essentially identical in the week corresponding to the peak of the outbreak, with a general shift to slightly higher Ct values (lower viral loads) in individuals without symptoms as the epidemic waned in Massachusetts. Over the entire time period, no clinically meaningful differences were observed overall, nor in each subgroup examined by age, sex, race, and ethnicity, despite some of the comparisons reaching statistical significance. By testing a large number of both residents and staff of nursing facilities, our study reports on viral load in a vulnerable subpopulation at risk for severe illness and death, as well as a younger and generally healthier staff population, with neither group exhibiting a meaningful overall difference in average Ct between individuals with and without symptoms at the time of testing.

As a group, individuals without symptoms at the time of testing had a distribution of viral loads similar to those measured in individuals with symptoms. There has been much discussion about potential heterogeneity in individuals who are labelled as asymptomatic at a single point in time, as they could be presymptomatic and will go on to develop symptoms in the future, post-symptomatic and are recovering, or durably asymptomatic and will never develop symptoms^1^. While this heterogeneity cannot be resolved without longitudinal follow-up, our point-prevalence study found that individuals without symptoms at the time of testing had viral loads that were similar to those from individuals with symptoms, with the viral loads being nearly identical during the peak of an epidemic when the time since infection acquisition is most similar between the two classes. These results suggest that the assay should be effective in detecting new infections in individuals without symptoms at the time of testing.

For individuals both with and without symptoms, viral loads detected on nasopharyngeal swabs varied by more than 250 million-fold, consistent with prior studies^18,19,20,21^ (Figure 1). The variation in viral load is much greater than seen for other factors that may affect infectivity (for example, the number of droplets expelled can vary by 100-fold across hosts^27^). Consistent with results for other respiratory illnesses^28^ and preliminary data for COVID-19 ^29 30^, it is plausible (but not proven) that infectivity of individuals with SARS-CoV-2 may be roughly proportional to viral load^11^. If so, a minority of individuals might account for the vast majority of transmission (Figure 2). For example, we calculated that 8-9% of individuals (either with or without symptoms) harbored 90% of the total viral load at the time of testing — which may partially explain the phenomenon of superspreading. Interestingly, our estimates are similar to recent inferences based on modeling of overdispersion in transmission, in which 10% of individuals may account for 80% of infections.^31^ (By contrast, the half of individuals with the lowest viral loads, those with Ct >26, carry only ∼0.01% of total viral load at the time of testing.)

The mean and distribution of Ct values for individuals with and without symptoms showed increasing differences over the 2 month duration of this study (Figure 3). The two groups were nearly identical in the first week in which symptom data were collected (Week 2 of the screening program, April 17-23), which corresponded to a few weeks after the statewide count of new cases began rising sharply (Figure S1). The mean viral load observed in individuals without symptoms then became lower in subsequent weeks, as the local prevalence subsided over the course of May. In fact, such a shift in the relationship between viral load and symptoms is expected based on local epidemic dynamics due to two factors. First, viral load changes over the course of an infection, increasing rapidly and then waning slowly over the course of weeks^8,32,33^, with the result that the viral loads observed at a given point in time will depend on the distribution of time since infection for the population studied. Second, in individuals who eventually develop them, symptoms are typically displayed within a limited time period early in the course of viral shedding, close to the peak of viral shedding^8^. As a consequence, even if the distribution of viral levels over time is identical between individuals who will and will not eventually develop symptoms, the set of individuals with symptoms at any given time will tend to be skewed toward more recent infections and thus higher viral levels compared to individuals without symptoms.

During the rapid initial growth phase of a local epidemic, and particularly in congregate settings where onset may be more synchronous, the skew in time-since-infection between those with and without symptoms might be expected to be modest because the vast majority of infections are recent. As a local epidemic stabilizes or declines, the skew would be expected to increase. Our data are consistent with this expectation, as the distribution of viral load in individuals without symptoms showed an increasing proportion of individuals with low viral loads over time, who are likely enriched for cases later in the course of infection.

Because the expected skew in time-since-infection is minimized in the rapid initial growth phase of a local epidemic, this period may provide a better representation of the prospective distribution of viral loads across individuals infected at roughly the same time. The fact that the distribution of viral load was initially nearly identical in individuals with and without symptoms suggests that whether an individual develops symptoms may not be primarily determined by viral load, but rather by other factors. However, longitudinal studies of both viral burden and symptoms are needed to clarify the relationship between viral load, symptoms, and clinical severity.

Notably, the majority of positive tests from both residents (70.8%) and staff (92.4%) came from individuals without symptoms at the time of testing. While each group of individuals may have been somewhat depleted for symptomatic COVID-19 (residents with severe symptoms may have been transferred to hospitals, while most symptomatic staff would likely have been required to stay home), these large percentages are consistent with smaller-scale cross-sectional studies in Iceland ^7^, Italy^34^, congregate facilities ^5,6,35,36,37^, labor and delivery wards in high-incidence cities ^2,3,4^, and the Diamond Princess cruise ship ^38,39,40^. However, the percent of truly asymptomatic infection remains an issue of much debate; smaller-scale longitudinal monitoring studies in a variety of settings vary considerably in their reported rates of symptom development among individuals who lacked symptoms at the time of a positive test ^36,38,39,40,41,37^. Nevertheless, modeling studies suggest that a substantial fraction of transmission occurs from people who are not symptomatic at the time, whether asymptomatic or pre-symptomatic^8^, which is reinforced by contact-tracing studies ^14,42,43^. Together, these findings underscore the need to expand beyond symptom-based screening as a sole tactic for detecting infected individuals and preventing transmission.

This study should be interpreted with certain caveats. First, without longitudinal follow-up, we cannot distinguish infected individuals who are permanently asymptomatic from those who are pre-symptomatic. However, both classes likely carry risk for transmitting the virus in the absence of symptoms ^8,14,42^ even while differing in their implications for contact tracing and for understanding the natural history of COVID-19^1^. Quantifying the viral burden in individuals without symptoms at the time of testing is thus an important step towards better understanding their transmission risk relative to symptomatic individuals. Second, with only a binary point-prevalence assessment of symptoms at the time of testing, we cannot draw any conclusions about the relationship between viral load and concurrent or future symptom severity in this population; however, the similarity in viral load distributions between individuals with and without symptoms suggests that viral load may not be the sole determinant of symptoms. These are important avenues for future study in longitudinal studies. Third, nursing home residents and staff may differ with respect to stages or disease severity from other populations, such as severely symptomatic individuals presenting to an acute setting for testing or requiring hospitalization^12^, or asymptomatic individuals in different settings. Nonetheless, these data represent Ct values for non-hospitalized individuals who did not seek acute testing, which represents the majority of COVID-19 cases and the vast majority of those at risk for ongoing transmission. Fourth, the widely-used approach of defining viral load based on RNA levels measured in specimens may not precisely reflect the number of live virions carried by an individual for several reasons. The assay may not reflect viral loads in other sites in the body and does not distinguish the genomic RNA of live virus from intact RNA from inactive or killed virus, which are thought to explain the long tail of low-level positive tests often seen during recovery^30^. In principle, the RNA level in a specimen could reflect both levels of full-length genomic RNA and subgenomic expression of the gene. (Expression has been reported to vary by ∼100-fold across the viral genome, with the N gene, targeted here, having higher levels^44^; however, this is much smaller than the >10^8^-fold differences in RNA levels observed across individuals.)

While our study found similar overall distributions in individuals with and without symptoms, our observation that the distributions began to diverge from the peak to later stages of the local epidemic suggests that substantial differences may be observed in other settings. We expect that the distributions seen in other settings will depend on both the selection of individuals for testing and the stage of the epidemic. Our study design — a cross-sectional study based on comprehensively sampling all individuals, independent of symptoms, at an early stage of the epidemic, when many cases are of similar age — is well-suited for understanding the prospective distribution of viral loads across infected individuals. At the opposite extreme, a cross-sectional study that tested inpatients who had been hospitalized for varying lengths of time due to severe COVID-19 symptoms would be expected to show a large difference in viral loads between individuals with and without symptoms, because those patients without symptoms at the time of testing would be entirely composed of later-stage recovering patients, in whom viral loads would be low. Similarly, if outpatients with a known exposure are tested either at the onset of symptoms for those who develop symptoms or at the end of a period of self-quarantine for “clearance” if they do not develop symptoms, the distributions of viral load in individuals with and without symptoms would be expected to differ substantially due to differences in average time since infection, not necessarily due to intrinsic differences in biology between the groups.

In summary, the majority of residents and the vast majority of staff who tested positive reported no symptoms at the time of sampling, and the viral loads in those with and without symptoms showed very similar distributions, particularly early in the study during the peak of the local epidemic. With testing of asymptomatic individuals under consideration in many settings, including contact tracing by public health departments and screening in workplaces or schools, a quantitative assessment of viral burden in individuals without symptoms is crucial to inform the viability of such screening strategies. While optimal implementation strategies and cost-effectiveness must be carefully considered, the finding of relatively similar viral load between infected individuals with and without symptoms at the time of testing builds confidence in the technical feasibility of identifying asymptomatic individuals harboring SARS-CoV-2 by standard RT-PCR assays.

## Data Availability

Underlying clinical data containing identifying information cannot be shared. De-identified data can be made available on request.

---

**Figure S1.**
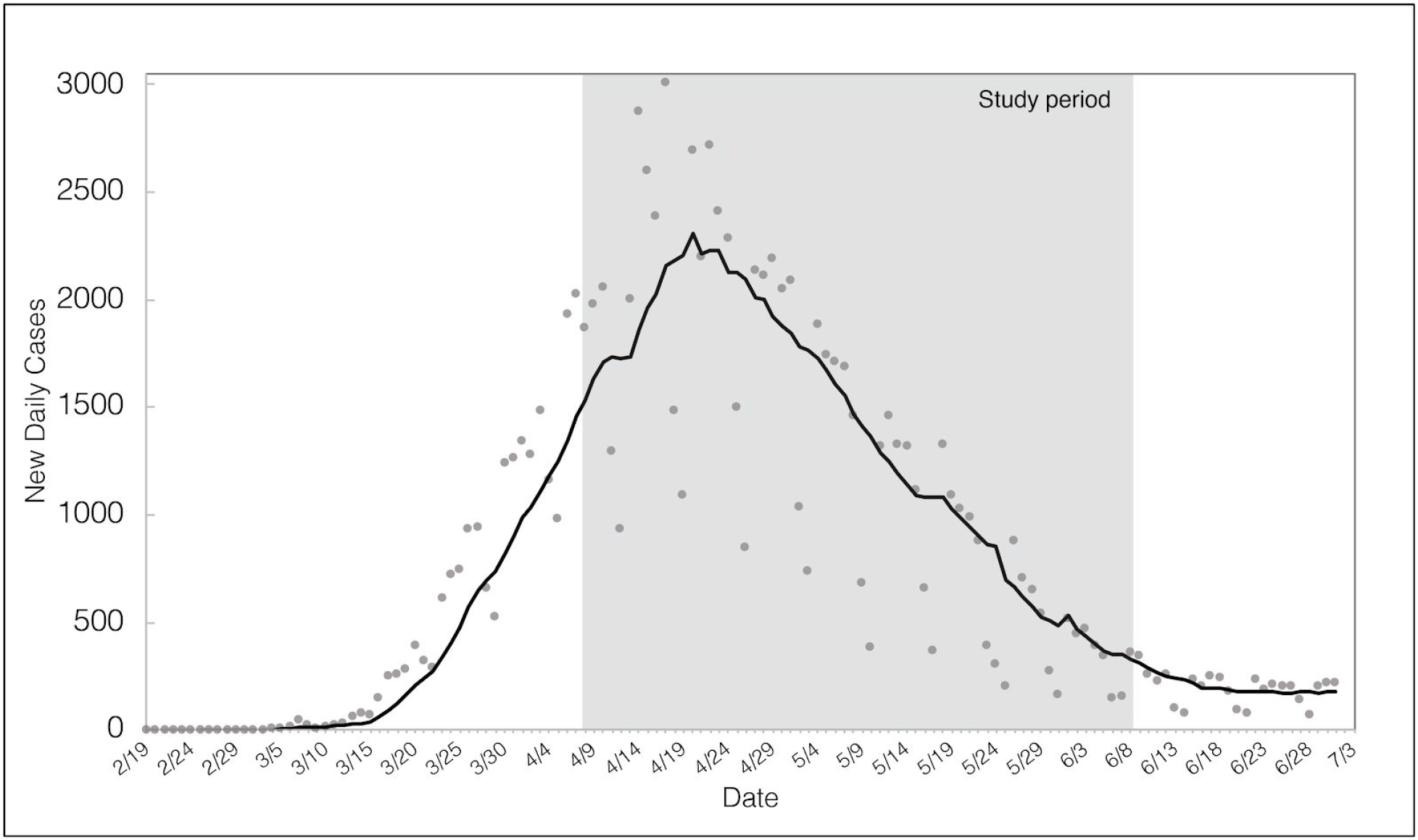
Time course of outbreak in MA. Daily confirmed cases (by date of sample collection) over time in Massachusetts, along with a 7-day moving average. Data taken from Massachusetts Department of Public Health COVID-19 Dashboard as of 7/14/2020 (see https://www.mass.gov/info-details/archive-of-covid-19-cases-in-massachusetts).

